# Influence of vitamin D supplementation on bone mineral content, bone turnover markers and fracture risk in South African schoolchildren: multicentre double-blind randomised placebo-controlled trial (ViDiKids)

**DOI:** 10.1101/2023.05.18.23290153

**Authors:** Keren Middelkoop, Lisa K Micklesfield, Neil Walker, Justine Stewart, Carmen Delport, David A Jolliffe, Amy E Mendham, Anna K Coussens, Averalda van Graan, James Nuttall, Jonathan C Y Tang, William D Fraser, Cyrus Cooper, Nicholas C Harvey, Richard L Hooper, Robert J Wilkinson, Linda-Gail Bekker, Adrian R Martineau

## Abstract

**BACKGROUND:** Randomised controlled trials (RCT) to determine the influence of vitamin D on bone mineral content (BMC) and fracture risk in children of Black African ancestry are lacking.

**METHODS:** We conducted a sub-study nested within a Phase 3 RCT of weekly oral supplementation with 10,000 IU vitamin D_3_ in HIV-uninfected Cape Town schoolchildren of Black African ancestry aged 6-11 years. Outcomes were BMC at the whole body less head (WBLH) and lumbar spine (LS) and serum concentrations of 25-hydroxyvitamin D_3_ (25[OH]D_3_), parathyroid hormone (PTH) and bone turnover markers. Incidence of fractures was an outcome of the main trial.

**FINDINGS:** 1682 children were enrolled in the main trial, of whom 450 also participated in the sub-study. Among sub-study participants, end-trial serum 25(OH)D_3_ concentrations were higher for participants allocated to vitamin D vs. placebo (adjusted mean difference [aMD] 39.9 nmol/L, 95% CI 36.1 to 43.6, P<0.001) and serum PTH concentrations were lower (aMD -0.55 pmol/L, 95% CI -0.94 to -0.17, P=0.005). However, no interarm differences were seen for WBLH BMC (aMD -8.0 g, 95% CI - 30.7 to 14.7) or LS BMC (aMD -0.3 g, 95% CI -1.3 to 0.8), or for serum concentrations of bone turnover markers (P≥0.28). In the main trial, allocation did not influence fracture risk (adjusted odds ratio 0.70, 95% CI 0.27 to 1.85, P=0.48).

**INTERPRETATION:** Weekly vitamin D supplementation elevated serum 25(OH)D_3_ concentrations and suppressed serum PTH concentrations in HIV-uninfected schoolchildren of Black African ancestry but did not influence BMC, bone turnover markers or fracture risk.

**FUNDING:** Medical Research Council

**RESEARCH IN CONTEXT:** *EVIDENCE BEFORE THIS STUDY:* We searched PubMed from inception to 31^st^ December 2022 for randomised controlled trials (RCT) evaluating effects of vitamin D supplementation on bone mineral content (BMC), bone mineral density (BMD) and fracture risk in HIV-uninfected schoolchildren. A meta-analysis of data from 884 participants in six RCT reported no statistically significant effects of vitamin D on total body BMC, hip BMD, or forearm BMD, but a trend towards a small positive effect on lumbar spine BMD. RCT investigating fracture outcomes in HIV-uninfected children were lacking, as were RCT investigating effects of vitamin D on bone outcomes in HIV-uninfected children of Black African ancestry.

*ADDED VALUE OF THIS STUDY:* This is the first RCT to investigate effects of vitamin D supplementation on BMC and fracture risk in HIV-uninfected schoolchildren of Black African ancestry. We found that weekly oral supplementation with 10,000 IU vitamin D_3_ for 3 years elevated serum 25(OH)D_3_ concentrations and suppressed serum PTH concentrations, but did not influence serum concentrations of bone turnover markers, BMC at the whole body less head or lumbar spine sites, or fracture risk.

*IMPLICATIONS OF ALL THE AVAILABLE EVIDENCE:* Taken together with null findings from another recenty-completed phase 3 RCT of weekly oral vitamin D supplementation conducted in Mongolian schoolchildren, our findings do not support a role for vitamin D supplementation to increase BMC or reduce fracture risk in primary schoolchildren.

## INTRODUCTION

Low bone mineral density (BMD) and related fractures cause a large and increasing global burden of disability-adjusted life years and mortality.^1^ Osteoporosis in adulthood has its origins in childhood, which is an important period for optimisation of bone mass.^2^ Vitamin D has long been recognised to play a key role in promoting bone mineralisation,^3^ and observational studies report associations between low circulating concentrations of 25-hydroxyvitamin D (25[OH]D) and increased fracture risk in children.^4^ Randomised controlled trials (RCT) of vitamin D supplementation to augment bone mineral content (BMC) in children and adolescents have yielded variable results,^5,6^ although there is some evidence to suggest that higher doses of vitamin D (e.g. >1000 IU/day or equivalent) may be more effective than lower doses^7-9^ and that intervention before or during the onset of adolescence may be associated with greater benefit in terms of BMC accumulation than later supplementation.^7,10,11^ However, despite evidence that relationships between vitamin D status, parathyroid hormone (PTH), BMD and fracture risk differ between children of White European vs. Black African ancestry,^12-14^ RCT to determine effects of vitamin D on BMC and bone turnover markers in African children are lacking.

In order to address this deficit, we performed a sub-study nested within the ViDiKids trial, a multicentre phase 3 RCT which investigated effects of weekly oral administration of 10,000 IU vitamin D_3_ for 3 years on the primary outcome of tuberculosis infection in a cohort of 1,682 schoolchildren aged 6-11 years living in a socio-economically disadvantaged peri-urban district of Cape Town, South Africa.^15^ Sub-study outcomes were BMC at the whole body less head and lumbar spine sites, and serum concentrations of 25[OH]D_3_, PTH, alkaline phosphatase (ALP), C-terminal telopeptide (CTX) and procollagen type 1 N propeptide (P1NP). The influence of vitamin D supplementation on incidence of fractures in the study population as a whole was also investigated.

## METHODS

### TRIAL DESIGN, SETTING, APPROVALS AND REGISTRATION

We conducted a multicentre phase 3 double-blind individually randomised placebo-controlled trial in 23 government schools in Cape Town, South Africa, as previously described.^15^ The primary outcome was acquisition of latent tuberculosis infection; the current manuscript reports effects of the intervention on pre-specified secondary outcomes relating to fracture incidence in all study participants, and BMC and serum concentrations of 25(OH)D_3_, adjusted calcium, PTH and markers of bone turnover in a subset of participants who additionally took part in a nested bone sub-study. The trial was sponsored by Queen Mary University of London, approved by the University of Cape Town Faculty of Health Sciences Human Research Ethics Committee (Ref: 796/2015) and the London School of Hygiene and Tropical Medicine Observational/Interventions Research Ethics Committee (Ref: 7450-2) and registered on the South African National Clinical Trials Register (DOH-27-0916-5527) and ClinicalTrials.gov (ref NCT02880982).

### PARTICIPANTS

Inclusion criteria for the main trial were enrolment in Grades 1-4 at a participating school; age 6 to 11 years at screening; and written informed assent / consent to participate in the main trial provided by children and their parent / legal guardian, respectively. Exclusion criteria for the main trial were a history of previous latent TB infection, active TB disease or any chronic illness other than asthma (including known or suspected HIV infection) prior to enrolment; use of any regular medication other than asthma medication; use of vitamin D supplements at a dose of more than 400 IU/day in the month before enrolment; plans to move away from study area within 3 years of enrolment; inability to swallow a placebo soft gel capsule with ease; and clinical evidence of rickets or a positive QFT-Plus assay result at screening. An additional inclusion criterion for the bone sub-study was enrolment in Grade 4 at a participating school.

### ENROLMENT

Parents or legal guardians were invited to provide written informed consent for their child to participate in the main trial during a home visit, unless their child was eligible for the bone sub-study, in which case they were invited to provide written informed consent for their child to participate in both the main trial and the bone sub-study until a total of 450 sub-study participants were randomised. If parents / legal guardians consented, they were asked to provide details of their child’s dietary intake of foods containing vitamin D and calcium in the previous month, which were captured on an electronic case report form (Fig. S1, Supplementary Material). Their children were then invited to provide written assent to participate in the main trial +/-the bone sub-study (if eligible) at a school-based visit. If they agreed, a clinically trained member of the study team screened them for symptoms and signs of rickets. For all participants, a blood sample was taken for a QFT-Plus assay and separation and storage of serum for determination of 25(OH)D concentrations as described below. For bone sub-study participants, additional blood was taken for determination of serum concentrations of calcium, albumin, PTH, total ALP, P1NP and CTX as described below. Participants were reviewed when baseline QFT-Plus results were available. Those with a positive QFT-Plus result were excluded from the trial and screened for active TB. Those with an indeterminate QFT-Plus result were excluded from the trial without screening for active TB. Those with a negative QFT-Plus result were deemed eligible to participate and underwent measurement of weight (using a digital floor scale, Charder Medical) and height (using a portable HM200P stadiometer, Charder Medical). Bone sub-study participants also underwent baseline dual energy x-ray absorptiometry (DXA) scanning as described below.

### RANDOMISATION AND BLINDING

Full details of randomisation and blinding procedures have been described previously^15^ and are presented in Supplementary Material. Briefly, eligible and assenting children whose parents consented to their participation in the trial were individually randomised to receive a weekly capsule containing vitamin D_3_ or placebo for three years, with a one-to-one allocation ratio and randomisation stratified by school of attendance. Treatment allocation was concealed from participants, care providers and all trial staff (including senior investigators and those assessing outcomes) until completion of the trial to maintain the double-blind.

### INTERVENTION

Study medication comprised a 3-year course of weekly soft gel capsules manufactured by the Tishcon Corporation (Westbury, NY, USA), containing either 0.25 mg (10,000 international units) cholecalciferol (vitamin D_3_) in olive oil (intervention arm) or olive oil without any vitamin D_3_ content (placebo arm). Active and placebo capsules had identical appearance and taste. Capsules were taken under direct observation of study staff during school termtime. During summer holidays (8 weeks), packs containing 8 doses of study medication were provided for administration by parents, together with a participant diary. Following shorter school holidays (≤4 weeks), and/or if participants missed one or more doses of study medication during term time, up to 4 ‘catch-up’ doses were administered at the first weekly visit attended following the missed dose(s). During the initial national lockdown for COVID-19 in South Africa (27^th^ March to 1^st^ May 2020), participants did not receive any study medication. During subsequent school closures due to COVID-19, two rounds of 8-week holiday packs were provided to participants, which were sufficient to cover their requirements until schools re-opened.

At weekly study visits during school terms, the study team captured data on adverse events and supervised the administration of study capsules. At 1-year, 2-year and 3-year follow-up, history of fractures in the previous year was captured using an electronic case report form (Fig. S2, Supplementary Material). At 3-year follow-up all participants were invited to provide a blood sample for QFT-Plus testing and separation and storage of serum for determination of 25(OH)D_3_ concentrations. Bone sub-study participants were invited to give extra blood for determination of end-study serum concentrations of calcium, albumin, PTH, total ALP, P1NP and CTX as described below, and to undergo repeat DXA scanning as at baseline.

### OUTCOMES

The primary outcome for the bone sub-study was BMC at the whole body less head and lumbar spine sites at 3-year follow-up. Secondary outcomes for the bone sub-study were serum concentrations of 25(OH)D_3_, adjusted calcium, PTH, ALP, CTX and P1NP. The primary outcome for the main trial was the QuantiFERON-TB Gold Plus result at the manufacturer-recommended 0.35 IU/mL threshold at the end of the study. Fracture incidence was a secondary outcome for the main trial.

### DXA

DXA scans were performed at the Sports Science Institute of South Africa, University of Cape Town, by a trained radiographer on one Hologic bone densitometer (Discovery-W®, Hologic, Bedford, MA, USA) using standard procedures, and analysed using Apex software (Version 13.4.1). Quality assurance checks were carried out prior to scanning and generated coefficients of variation <0.5%. Whole body less head and lumbar spine scans were performed to measure BMC with and without volumetric correction and correction for bone area, height and weight as described elsewhere.^16^

### LABORATORY ASSESSMENTS

Biochemical analyses were performed at the Bioanalytical Facility, University of East Anglia (Norwich, UK) according to manufacturers’ instructions and under Good Clinical and Laboratory Practice conditions. Serum concentrations of 25(OH)D_3_ were measured using liquid chromatography tandem mass spectrometry (LC-MS/MS) as previously described.^17^ 25(OH)D_3_ was calibrated using standard reference material SRM972a from the National Institute of Science and Technology (NIST), and the assay showed linearity between 0 and 200 nmol/L. The inter/intra-assay coefficient of variation (CV) across the assay range was ≤9%, and the lower limit of quantification was 0.1 nmol/L. The assay showed <6% accuracy bias against NIST reference method on the vitamin D external quality assessment (DEQAS) scheme (http://www.deqas.org/; accessed on 30^th^ November 2022). Serum concentrations of total calcium, albumin and creatinine were measured by spectrophotometric methods on the Cobas c501 platform (Roche Diagnostics, Penzberg, Germany) according to the manufacturer’s instructions. The inter-assay CV for total calcium and albumin were ≤ 2.1% across the assay working ranges of 0.2 to 7.5 mmol/L and 2 to 60 g/L. Albumin-adjusted calcium was calculated as total calcium (mmol/l) + 0.02 × (40 – albumin [g/l]). Serum ALP concentrations were measured by colourimetric assay on the Cobas e501 platform (Roche): the inter-assay CV across the assay working range of 5-1200 U/L was ≤2.4%. Serum concentrations of CTX, PINP, PTH and total ALP were measured using electrochemiluminesence immunoassays (ECLIA) performed on the Cobas e601 platform (Roche). The inter-assay CV for CTX was ≤3% between 0.2 and 1.5 μg/L with a sensitivity of 0.01 μg/L. The inter-assay CV for P1NP was ≤3% between 20-600 μg/L with a sensitivity of 8 μg/L. The inter-assay CV for PTH was ≤3.8% between 0.127-530 pmoL/L. QFT-Plus assays were performed by the Bio Analytical Research Corporation South Africa (Johannesburg, South Africa) according to the manufacturer’s instructions.

### SAMPLE SIZE

Sample size for the main trial was predicated on power to detect an effect of the intervention on the primary outcome (the proportion of children with a positive QFT-Plus assay result at 3-year follow-up), as previously described.^15^ The bone sub-study was powered to detect a clinically significant effect of vitamin D on BMC: assuming 29% loss to follow-up at 3 years, we calculated that enrolment of 450 participants would provide 88% power to detect a difference of 0.35 standard deviations between arms for mean BMC at either site investigated at the 5% significance level.

### STATISTICAL ANALYSES

Statistical analyses were performed using Stata software (Version 17.0; StataCorp, College Station, Texas, United States) according to intention to treat. Effects of allocation to vitamin D vs. placebo on BMC and other continuous outcomes were estimated using mixed-effects linear regression with adjustment for baseline value and a random effect of school of attendance, with results reported as treatment differences with 95% confidence intervals. Pre-specified sub-group analyses were conducted to determine whether the effect of vitamin D supplementation was modified by sex (male vs. female), baseline deseasonalised 25(OH)D_3_ concentration, calculated using a sinusoidal model as previously described^18^ (<75 vs. ≥75 nmol/L) and calcium intake, calculated as described in Supplementary Material, Table S1 (< vs. ≥median value of 466 mg/day). These were performed by repeating efficacy analyses with the inclusion of an interaction term between allocation (to vitamin D vs. placebo) and each posited effect-modifier with presentation of the P-value associated with this interaction term. Given the number of potential effect modifiers and secondary outcome measures these analyses are considered exploratory. Effects of treatment on the proportion of participants reporting one or more fractures were estimated by fitting allocation (vitamin D vs. placebo) as the sole fixed effect in a mixed effects logistic regression model with a random effect for repeated assessments of each individual participant (in years 1, 2 and 3 of follow-up) and a random effect of school of attendance. Results are reported as odds ratios with 95% confidence intervals. Interim safety assessments, where Independent Data Monitoring Committee (IDMC) members reviewed accumulating serious adverse event data, were performed at 6-monthly intervals. At each review the IDMC recommended continuation of the trial. No interim efficacy analysis was performed.

## RESULTS

### PARTICIPANTS

Of 2852 children screened for eligibility from March 2017 to March 2019, 2271 underwent QFT-Plus testing. 1682 (74.1%) of these tested negative and were randomly assigned to receive vitamin D_3_ (829 participants) or placebo (853 participants) as previously described.^15^ 450/1682 (26.8%) participants in the main trial also participated in the bone sub-study, of whom 228 vs. 222 participants were allocated to the vitamin D vs. placebo arms, respectively (Fig. 1). Table 1 presents baseline characteristics of children in the main trial and in the bone sub-study, overall and by study arm. Mean age was higher among participants in the sub-study vs. all those in the main trial (10.1 vs. 8.9 years, respectively), reflecting the fact that participation in the sub-study was restricted to children enrolled in Grade 4. Baseline characteristics were otherwise well balanced for all participants in the main trial vs. those who additionally participated in the sub-study: 52.4% vs. 52.0% were female and mean serum 25[OH]D_3_ concentrations were 71.2 nmol/L vs. 70.0 nmol/L, respectively. Within the main trial and the sub-study, baseline characteristics of those randomised to vitamin D vs. placebo were also well balanced.

**Figure 1:**
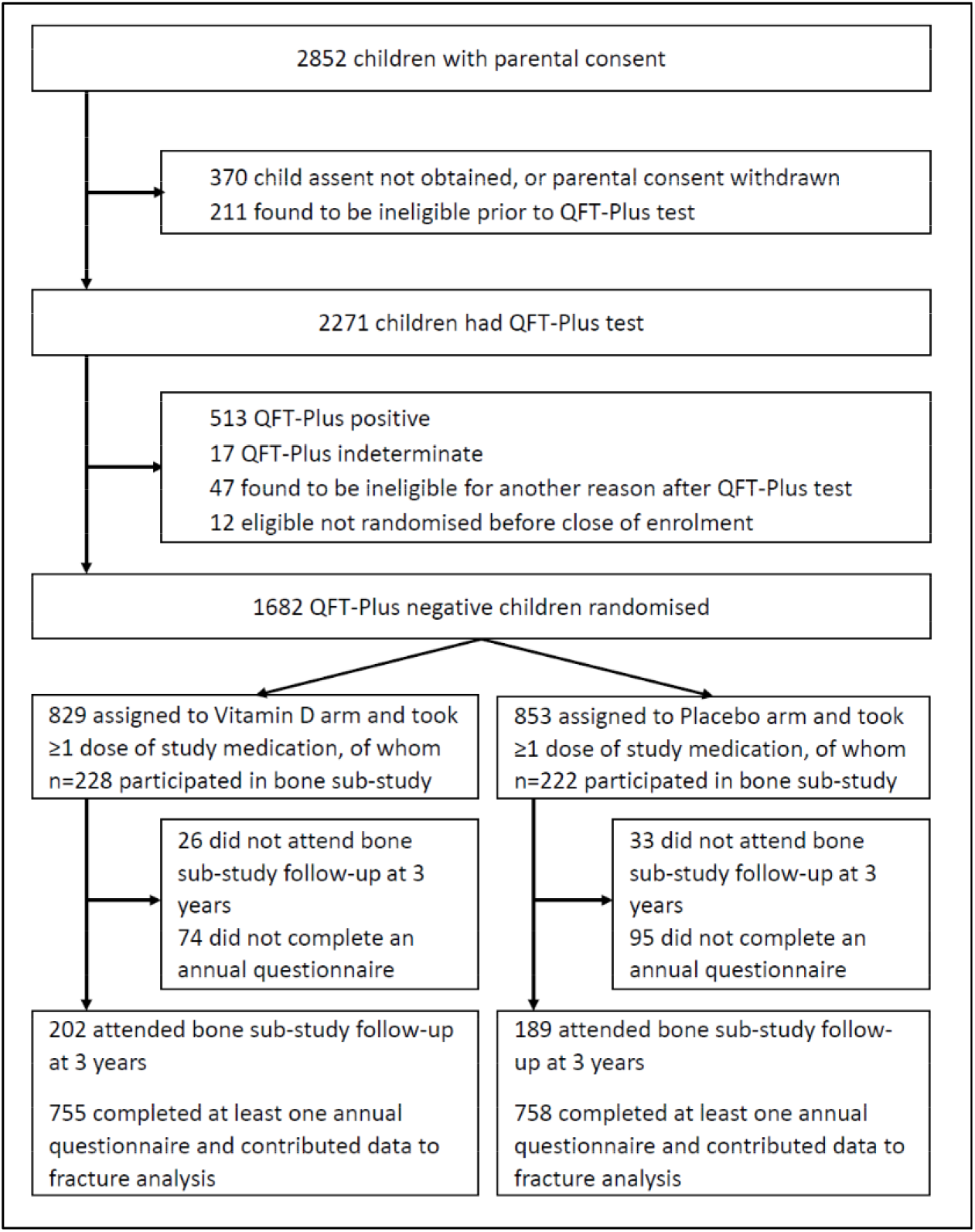
Participant flow

**Table 1:**
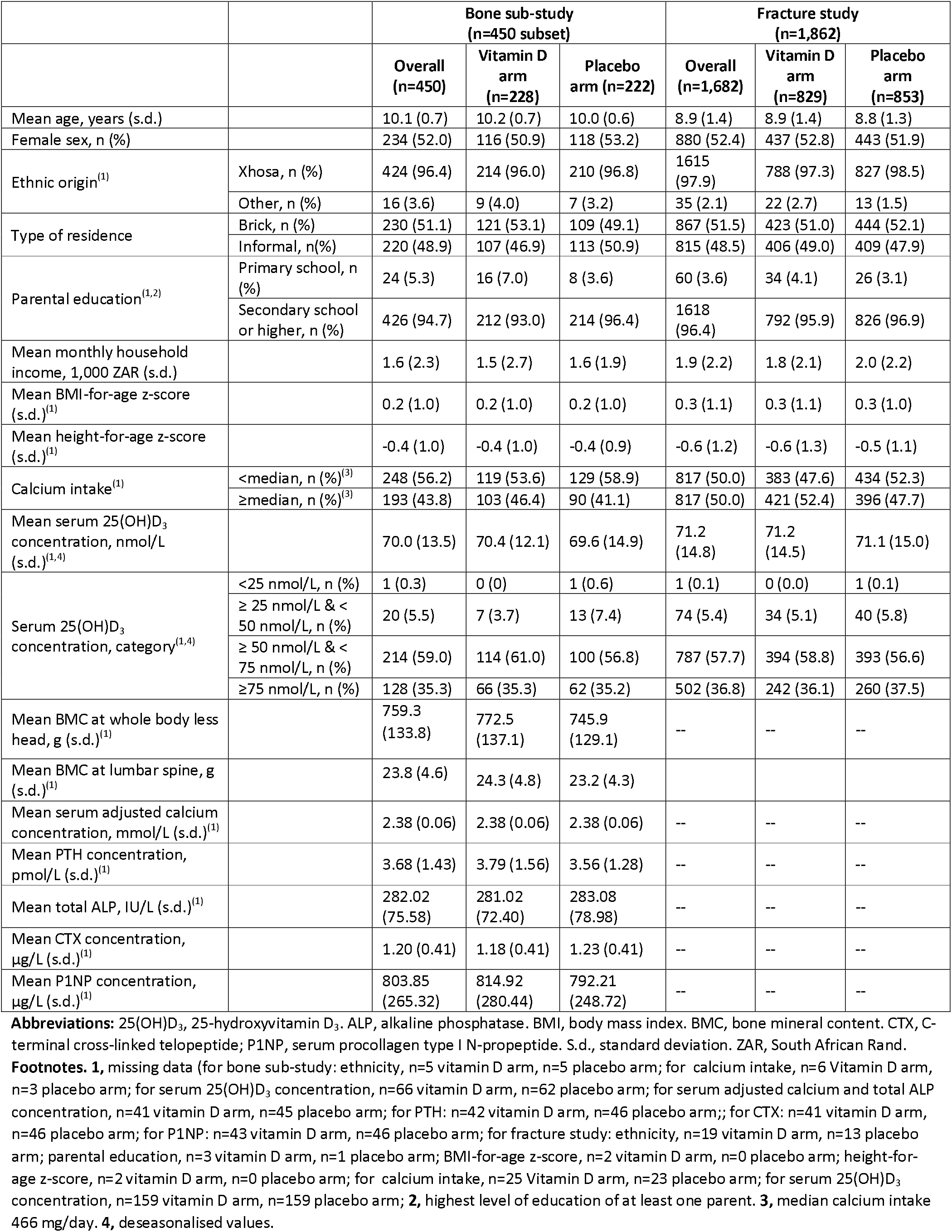
Participants’ baseline characteristics by allocation: bone sub-study and fracture study

|  |  | Bone sub-study<br>(n=450 subset) |  |  | Fracture study<br>(n=1,862) |  |  |
| --- | --- | --- | --- | --- | --- | --- | --- |
|  |  | Overall<br>(n=450) | Vitamin D<br>arm<br>(n=228) | Placebo<br>arm (n=222) | Overall<br>(n=1,682) | Vitamin D<br>arm<br>(n=829) | Placebo<br>arm<br>(n=853) |
| Mean age, years (s.d.) |  | 10.1 (0.7) | 10.2 (0.7) | 10.0 (0.6) | 8.9 (1.4) | 8.9 (1.4) | 8.8 (1.3) |
| Female sex, n (%) |  | 234 (52.0) | 116 (50.9) | 118 (53.2) | 880 (52.4) | 437 (52.8) | 443 (51.9) |
| Ethnic origin<sup>(1)</sup> | Xhosa, n (%) | 424 (96.4) | 214 (96.0) | 210 (96.8) | 1615<br>(97.9) | 788 (97.3) | 827 (98.5) |
|  | Other, n (%) | 16 (3.6) | 9 (4.0) | 7 (3.2) | 35 (2.1) | 22 (2.7) | 13 (1.5) |
| Type of residence | Brick, n (%) | 230 (51.1) | 121 (53.1) | 109 (49.1) | 867 (51.5) | 423 (51.0) | 444 (52.1) |
|  | Informal, n(%) | 220 (48.9) | 107 (46.9) | 113 (50.9) | 815 (48.5) | 406 (49.0) | 409 (47.9) |
| Parental education<sup>(1,2)</sup> | Primary school, n (%) | 24 (5.3) | 16 (7.0) | 8 (3.6) | 60 (3.6) | 34 (4.1) | 26 (3.1) |
|  | Secondary school or higher, n (%) | 426 (94.7) | 212 (93.0) | 214 (96.4) | 1618<br>(96.4) | 792 (95.9) | 826 (96.9) |
| Mean monthly household income, 1,000 ZAR (s.d.) |  | 1.6 (2.3) | 1.5 (2.7) | 1.6 (1.9) | 1.9 (2.2) | 1.8 (2.1) | 2.0 (2.2) |
| Mean BMI-for-age z-score (s.d.)<sup>(1)</sup> |  | 0.2 (1.0) | 0.2 (1.0) | 0.2 (1.0) | 0.3 (1.1) | 0.3 (1.1) | 0.3 (1.0) |
| Mean height-for-age z-score (s.d.)<sup>(1)</sup> |  | -0.4 (1.0) | -0.4 (1.0) | -0.4 (0.9) | -0.6 (1.2) | -0.6 (1.3) | -0.5 (1.1) |
| Calcium intake<sup>(1)</sup> | <median, n (%)<sup>(3)</sup> | 248 (56.2) | 119 (53.6) | 129 (58.9) | 817 (50.0) | 383 (47.6) | 434 (52.3) |
|  | ≥median, n (%)<sup>(3)</sup> | 193 (43.8) | 103 (46.4) | 90 (41.1) | 817 (50.0) | 421 (52.4) | 396 (47.7) |
| Mean serum 25(OH)D<sub>3</sub> concentration, nmol/L (s.d.)<sup>(1,4)</sup> |  | 70.0 (13.5) | 70.4 (12.1) | 69.6 (14.9) | 71.2<br>(14.8) | 71.2<br>(14.5) | 71.1 (15.0) |
| Serum 25(OH)D<sub>3</sub> concentration, category<sup>(1,4)</sup> | <25 nmol/L, n (%) | 1 (0.3) | 0 (0) | 1 (0.6) | 1 (0.1) | 0 (0.0) | 1 (0.1) |
|  | ≥ 25 nmol/L & < 50 nmol/L, n (%) | 20 (5.5) | 7 (3.7) | 13 (7.4) | 74 (5.4) | 34 (5.1) | 40 (5.8) |
|  | ≥ 50 nmol/L & < 75 nmol/L, n (%) | 214 (59.0) | 114 (61.0) | 100 (56.8) | 787 (57.7) | 394 (58.8) | 393 (56.6) |
|  | ≥75 nmol/L, n (%) | 128 (35.3) | 66 (35.3) | 62 (35.2) | 502 (36.8) | 242 (36.1) | 260 (37.5) |
| Mean BMC at whole body less head, g (s.d.)<sup>(1)</sup> |  | 759.3<br>(133.8) | 772.5<br>(137.1) | 745.9<br>(129.1) | -- | -- | -- |
| Mean BMC at lumbar spine, g (s.d.)<sup>(1)</sup> |  | 23.8 (4.6) | 24.3 (4.8) | 23.2 (4.3) |  |  |  |
| Mean serum adjusted calcium concentration, mmol/L (s.d.)<sup>(1)</sup> |  | 2.38 (0.06) | 2.38 (0.06) | 2.38 (0.06) | -- | -- | -- |
| Mean PTH concentration, pmol/L (s.d.)<sup>(1)</sup> |  | 3.68 (1.43) | 3.79 (1.56) | 3.56 (1.28) | -- | -- | -- |
| Mean total ALP, IU/L (s.d.)<sup>(1)</sup> |  | 282.02<br>(75.58) | 281.02<br>(72.40) | 283.08<br>(78.98) | -- | -- | -- |
| Mean CTX concentration, µg/L (s.d.)<sup>(1)</sup> |  | 1.20 (0.41) | 1.18 (0.41) | 1.23 (0.41) | -- | -- | -- |
| Mean P1NP concentration, µg/L (s.d.)<sup>(1)</sup> |  | 803.85<br>(265.32) | 814.92<br>(280.44) | 792.21<br>(248.72) | -- | -- | -- |

**Abbreviations:** 25(OH)D<sub>3</sub>, 25-hydroxyvitamin D<sub>3</sub>. ALP, alkaline phosphatase. BMI, body mass index. BMC, bone mineral content. CTX, C-terminal cross-linked telopeptide; P1NP, serum procollagen type I N-propeptide. S.d., standard deviation. ZAR, South African Rand.
**Footnotes.** 1, missing data (for bone sub-study: ethnicity, n=5 vitamin D arm, n=5 placebo arm; for calcium intake, n=6 Vitamin D arm, n=3 placebo arm; for serum 25(OH)D<sub>3</sub> concentration, n=66 vitamin D arm, n=62 placebo arm; for serum adjusted calcium and total ALP concentration, n=41 vitamin D arm, n=45 placebo arm; for PTH: n=42 vitamin D arm, n=46 placebo arm;; for CTX: n=41 vitamin D arm, n=46 placebo arm; for P1NP: n=43 vitamin D arm, n=46 placebo arm; for fracture study: ethnicity, n=19 vitamin D arm, n=13 placebo arm; parental education, n=3 vitamin D arm, n=1 placebo arm; BMI-for-age z-score, n=2 vitamin D arm, n=0 placebo arm; height-for-age z-score, n=2 vitamin D arm, n=0 placebo arm; for calcium intake, n=25 Vitamin D arm, n=23 placebo arm; for serum 25(OH)D<sub>3</sub> concentration, n=159 vitamin D arm, n=159 placebo arm; 2, highest level of education of at least one parent. 3, median calcium intake 466 mg/day. 4, deseasonalised values.

The median duration of follow-up was 3.16 years (interquartile range, 2.83 to 3.38 years) and was not different between the two study arms. For the main trial, mean serum 25(OH)D_3_ concentrations at 3-year follow-up were higher among children randomised to receive vitamin D vs. placebo (104.3 vs. 64.7 nmol/L, respectively; mean difference 39.7 nmol/L, 95% CI for difference 37.6 to 41.9 nmol/L).

### BONE MINERAL CONTENT

Table 2 presents values for mean BMC at the whole body less head and lumbar spine sites at 3-year follow-up by allocation. No difference in either outcome was seen between participants randomised to vitamin D vs. placebo overall (for whole body less head: 1112.9 vs. 1071.5 g respectively, adjusted mean difference [aMD] - 8.0, 95% CI -30.7 to 14.7, P=0.49; for lumbar spine: 36.2 vs. 34.2 respectively, Amd -0.3, 95% CI -1.3 to 0.8, P=0.65). Sub-group analysis by sex, baseline 25(OH)D_3_ concentration and calcium intake did not reveal evidence of effect modification by any of these factors (P values for interaction ≥0.11)). Overall results were also null when statistical analyses were conducted with volumetric correction and correction for bone area, height and weight (Table S2, Supplementary Material).

**Table 2:** Uncorrected^(1)^ end-trial bone mineral content at the whole body less head and lumbar spine sites by allocation: overall and by sub-groups.

|  |  | Vitamin D arm:<br>mean value, g (s.d.)<br>[N] | Placebo arm: mean<br>value, g (s.d.) [N] | Adjusted mean<br>difference (95% CI)<sup>(2)</sup> | P | P for<br>interaction |
| --- | --- | --- | --- | --- | --- | --- |
| Whole body less head |  |  |  |  |  |  |
| Overall |  | 1112.9 (255.3) [202] | 1071.5 (221.9) [189] | -8.0 (-30.7 to 14.7) | 0.49 | -- |
| By sex | Male | 1102.3 (268.5) [97] | 1031.4 (225.4) [89] | 1.9 (-32.0 to 35.7) | 0.91 | 0.11 |
|  | Female | 1122.7 (243.4) [105] | 1107.2 (213.5) [100] | -20.8 (-46.8 to 5.3) | 0.12 |  |
| By baseline 25(OH)D<sub>3</sub><br>concentration<sup>(3)</sup> | <75<br>nmol/L | 1137.0 (270.8) [103] | 1099.1 (224.2) [96] | -16.5 (-47.5 to 14.5) | 0.30 | 0.76 |
|  | ≥75<br>nmol/L | 1119.3 (269.6) [60] | 1058.4 (215.9) [54] | -8.4 (-50.5 to 33.7) | 0.70 |  |
| By calcium intake | <median<sup>(4)</sup> | 1111.3 (249.5) [94] | 1057.8 (212.9) [97] | 5.2 (-27.6 to 38.1) | 0.75 | 0.23 |
|  | ≥median<sup>(4)</sup> | 1120.7 (263.6) [102] | 1094.0 (228.9) [89] | -19.3 (-50.9 to 12.3) | 0.23 |  |
| Lumbar spine |  |  |  |  |  |  |
| Overall |  | 36.2 (10.1) [202] | 34.2 (8.0) [189] | -0.3 (-1.3 to 0.8) | 0.65 | -- |
| By sex | Male | 33.2 (9.7) [97] | 31.1 (6.8) [89] | 0.2 (-1.2 to 1.5) | 0.81 | 0.28 |
|  | Female | 39.0 (9.7) [105] | 37.0 (8.0) [100] | -0.5 (-1.7 to 0.7) | 0.40 |  |
| By baseline 25(OH)D<sub>3</sub><br>concentration<sup>(3)</sup> | <75<br>nmol/L | 37.5 (10.7) [103] | 35.4 (8.4) [96] | -0.2 (-1.6 to 1.3) | 0.82 | 0.82 |
|  | ≥75<br>nmol/L | 36.2 (10.5) [60] | 33.1 (6.9) [54] | -0.32(-2.4 to 1.8) | 0.77 |  |
| By calcium intake | <median | 35.5 (9.9) [94] | 33.9 (7.8) [97] | 0.1 (-1.4 to 1.6) | 0.93 | 0.54 |
|  | ≥median | 36.9 (10.2) [102] | 34.8 (8.2) [89] | -0.6 (-2.1 to 0.9) | 0.45 |  |

**Abbreviations:** 25(OH)D<sub>3</sub>, 25-hydroxyvitamin D<sub>3</sub>. BMC, bone mineral content. CI, confidence interval. S.d., standard deviation. N, number.
**Footnotes.** **1**, i.e. without volumetric correction or correction for bone area, height and weight. **2**, adjusted for baseline value and school of attendance. **3**, deseasonalised values. **4**, median calcium intake 466 mg/day.

### BIOCHEMICAL OUTCOMES

Table 3 presents mean values for serum concentrations of 25(OH)D_3_, adjusted calcium, PTH and bone turnover markers at 3-year follow-up in bone sub-study participants by allocation. In analyses of the sub-study population as a whole, mean serum 25(OH)D_3_ concentration at 3 years was higher among participants allocated to vitamin D vs. placebo (aMD 39.9 nmol/L, 95% CI for difference 36.1 to 43.6 nmol/L, P<0.001), and mean serum PTH concentration was lower (aMD -0.55 pmol/L, 95% CI -0.94 to -0.17, P=0.005); however, no inter-arm differences in end-study serum concentrations of adjusted calcium, alkaline phosphatase, CTX or P1NP were seen. Sub-group analyses indicated that effects of vitamin D were modified by baseline vitamin D status for the outcome of serum 25(OH)D_3_ concentration (P for interaction 0.04); by calcium intake for the outcome of ALP concentration (P for interaction 0.02); and by sex for the outcomes of serum CTX and P1NP concentrations (P values for interaction 0.03 and 0.049, respectively). P values for interaction were ≥0.10 for all other sub-group analyses of biochemical outcomes.

**Table 3.**
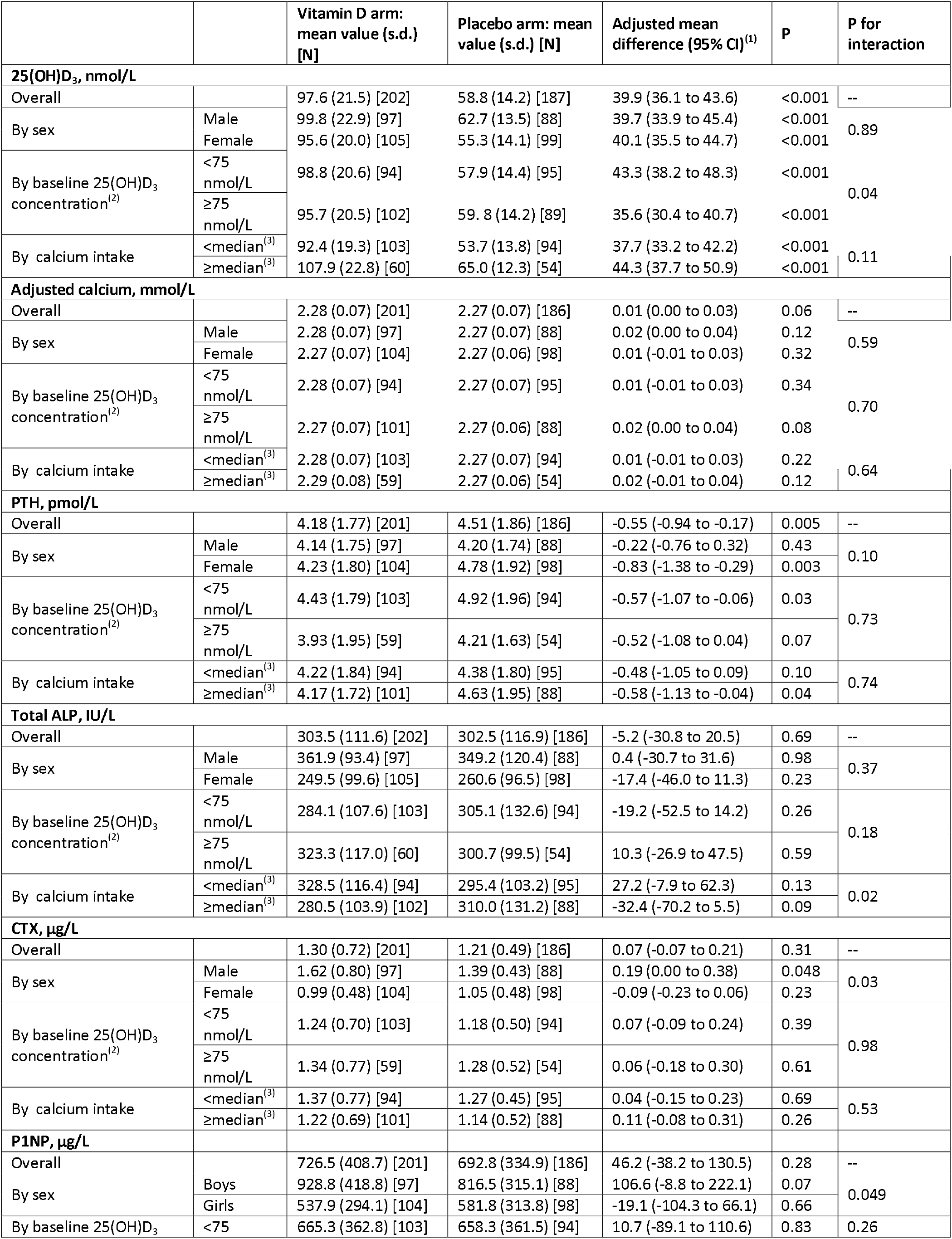
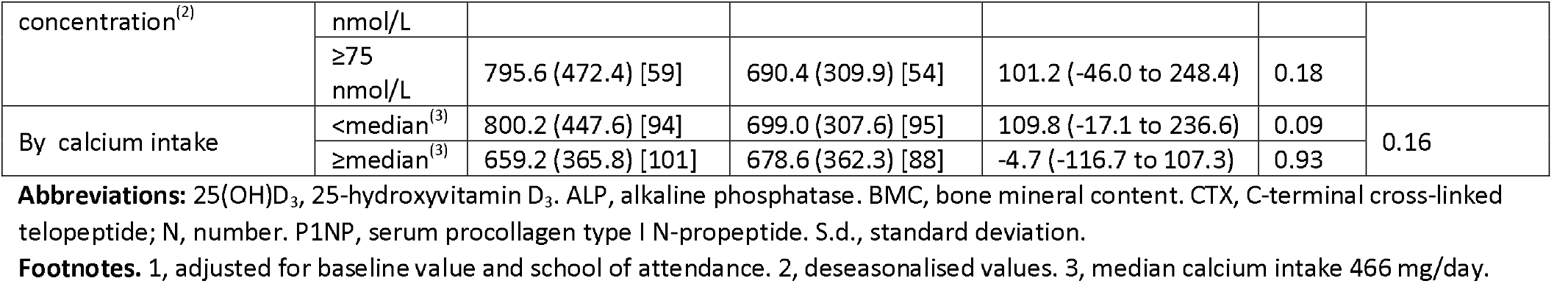
Serum concentrations of 25(OH)D_3_, adjusted calcium, PTH and bone turnover markers at 3-year follow-up by allocation: bone sub-study, overall and by sub-group

|  |  | Vitamin D arm:<br>mean value (s.d.)<br>[N] | Placebo arm: mean<br>value (s.d.) [N] | Adjusted mean<br>difference (95% CI)<sup>(1)</sup> | P | P for<br>interaction |
| --- | --- | --- | --- | --- | --- | --- |
| 25(OH)D<sub>3</sub>, nmol/L |  |  |  |  |  |  |
| Overall |  | 97.6 (21.5) [202] | 58.8 (14.2) [187] | 39.9 (36.1 to 43.6) | <0.001 | -- |
| By sex | Male | 99.8 (22.9) [97] | 62.7 (13.5) [88] | 39.7 (33.9 to 45.4) | <0.001 | 0.89 |
|  | Female | 95.6 (20.0) [105] | 55.3 (14.1) [99] | 40.1 (35.5 to 44.7) | <0.001 |  |
| By baseline 25(OH)D<sub>3</sub><br>concentration<sup>(2)</sup> | <75<br>nmol/L | 98.8 (20.6) [94] | 57.9 (14.4) [95] | 43.3 (38.2 to 48.3) | <0.001 | 0.04 |
|  | ≥75<br>nmol/L | 95.7 (20.5) [102] | 59.8 (14.2) [89] | 35.6 (30.4 to 40.7) | <0.001 |  |
| By calcium intake | <median<sup>(3)</sup> | 92.4 (19.3) [103] | 53.7 (13.8) [94] | 37.7 (33.2 to 42.2) | <0.001 | 0.11 |
|  | ≥median<sup>(3)</sup> | 107.9 (22.8) [60] | 65.0 (12.3) [54] | 44.3 (37.7 to 50.9) | <0.001 |  |
| Adjusted calcium, mmol/L |  |  |  |  |  |  |
| Overall |  | 2.28 (0.07) [201] | 2.27 (0.07) [186] | 0.01 (0.00 to 0.03) | 0.06 | -- |
| By sex | Male | 2.28 (0.07) [97] | 2.27 (0.07) [88] | 0.02 (0.00 to 0.04) | 0.12 | 0.59 |
|  | Female | 2.27 (0.07) [104] | 2.27 (0.06) [98] | 0.01 (-0.01 to 0.03) | 0.32 |  |
| By baseline 25(OH)D<sub>3</sub><br>concentration<sup>(2)</sup> | <75<br>nmol/L | 2.28 (0.07) [94] | 2.27 (0.07) [95] | 0.01 (-0.01 to 0.03) | 0.34 | 0.70 |
|  | ≥75<br>nmol/L | 2.27 (0.07) [101] | 2.27 (0.06) [88] | 0.02 (0.00 to 0.04) | 0.08 |  |
| By calcium intake | <median<sup>(3)</sup> | 2.28 (0.07) [103] | 2.27 (0.07) [94] | 0.01 (-0.01 to 0.03) | 0.22 | 0.64 |
|  | ≥median<sup>(3)</sup> | 2.29 (0.08) [59] | 2.27 (0.06) [54] | 0.02 (-0.01 to 0.04) | 0.12 |  |
| PTH, pmol/L |  |  |  |  |  |  |
| Overall |  | 4.18 (1.77) [201] | 4.51 (1.86) [186] | -0.55 (-0.94 to -0.17) | 0.005 | -- |
| By sex | Male | 4.14 (1.75) [97] | 4.20 (1.74) [88] | -0.22 (-0.76 to 0.32) | 0.43 | 0.10 |
|  | Female | 4.23 (1.80) [104] | 4.78 (1.92) [98] | -0.83 (-1.38 to -0.29) | 0.003 |  |
| By baseline 25(OH)D<sub>3</sub><br>concentration<sup>(2)</sup> | <75<br>nmol/L | 4.43 (1.79) [103] | 4.92 (1.96) [94] | -0.57 (-1.07 to -0.06) | 0.03 | 0.73 |
|  | ≥75<br>nmol/L | 3.93 (1.95) [59] | 4.21 (1.63) [54] | -0.52 (-1.08 to 0.04) | 0.07 |  |
| By calcium intake | <median<sup>(3)</sup> | 4.22 (1.84) [94] | 4.38 (1.80) [95] | -0.48 (-1.05 to 0.09) | 0.10 | 0.74 |
|  | ≥median<sup>(3)</sup> | 4.17 (1.72) [101] | 4.63 (1.95) [88] | -0.58 (-1.13 to -0.04) | 0.04 |  |
| Total ALP, IU/L |  |  |  |  |  |  |
| Overall |  | 303.5 (111.6) [202] | 302.5 (116.9) [186] | -5.2 (-30.8 to 20.5) | 0.69 | -- |
| By sex | Male | 361.9 (93.4) [97] | 349.2 (120.4) [88] | 0.4 (-30.7 to 31.6) | 0.98 | 0.37 |
|  | Female | 249.5 (99.6) [105] | 260.6 (96.5) [98] | -17.4 (-46.0 to 11.3) | 0.23 |  |
| By baseline 25(OH)D<sub>3</sub><br>concentration<sup>(2)</sup> | <75<br>nmol/L | 284.1 (107.6) [103] | 305.1 (132.6) [94] | -19.2 (-52.5 to 14.2) | 0.26 | 0.18 |
|  | ≥75<br>nmol/L | 323.3 (117.0) [60] | 300.7 (99.5) [54] | 10.3 (-26.9 to 47.5) | 0.59 |  |
| By calcium intake | <median<sup>(3)</sup> | 328.5 (116.4) [94] | 295.4 (103.2) [95] | 27.2 (-7.9 to 62.3) | 0.13 | 0.02 |
|  | ≥median<sup>(3)</sup> | 280.5 (103.9) [102] | 310.0 (131.2) [88] | -32.4 (-70.2 to 5.5) | 0.09 |  |
| CTX, µg/L |  |  |  |  |  |  |
| Overall |  | 1.30 (0.72) [201] | 1.21 (0.49) [186] | 0.07 (-0.07 to 0.21) | 0.31 | -- |
| By sex | Male | 1.62 (0.80) [97] | 1.39 (0.43) [88] | 0.19 (0.00 to 0.38) | 0.048 | 0.03 |
|  | Female | 0.99 (0.48) [104] | 1.05 (0.48) [98] | -0.09 (-0.23 to 0.06) | 0.23 |  |
| By baseline 25(OH)D<sub>3</sub><br>concentration<sup>(2)</sup> | <75<br>nmol/L | 1.24 (0.70) [103] | 1.18 (0.50) [94] | 0.07 (-0.09 to 0.24) | 0.39 | 0.98 |
|  | ≥75<br>nmol/L | 1.34 (0.77) [59] | 1.28 (0.52) [54] | 0.06 (-0.18 to 0.30) | 0.61 |  |
| By calcium intake | <median<sup>(3)</sup> | 1.37 (0.77) [94] | 1.27 (0.45) [95] | 0.04 (-0.15 to 0.23) | 0.69 | 0.53 |
|  | ≥median<sup>(3)</sup> | 1.22 (0.69) [101] | 1.14 (0.52) [88] | 0.11 (-0.08 to 0.31) | 0.26 |  |
| P1NP, µg/L |  |  |  |  |  |  |
| Overall |  | 726.5 (408.7) [201] | 692.8 (334.9) [186] | 46.2 (-38.2 to 130.5) | 0.28 | -- |
| By sex | Boys | 928.8 (418.8) [97] | 816.5 (315.1) [88] | 106.6 (-8.8 to 222.1) | 0.07 | 0.049 |
|  | Girls | 537.9 (294.1) [104] | 581.8 (313.8) [98] | -19.1 (-104.3 to 66.1) | 0.66 |  |
| By baseline 25(OH)D<sub>3</sub> | <75 | 665.3 (362.8) [103] | 658.3 (361.5) [94] | 10.7 (-89.1 to 110.6) | 0.83 | 0.26 |
| concentration<sup>(2)</sup> | nmol/L |  |  |  |  |  |
|  | ≥75 nmol/L | 795.6 (472.4) [59] | 690.4 (309.9) [54] | 101.2 (-46.0 to 248.4) | 0.18 |  |
| By calcium intake | <median<sup>(3)</sup> | 800.2 (447.6) [94] | 699.0 (307.6) [95] | 109.8 (-17.1 to 236.6) | 0.09 | 0.16 |
|  | ≥median<sup>(3)</sup> | 659.2 (365.8) [101] | 678.6 (362.3) [88] | -4.7 (-116.7 to 107.3) | 0.93 |  |

**Abbreviations:** 25(OH)D<sub>3</sub>, 25-hydroxyvitamin D<sub>3</sub>. ALP, alkaline phosphatase. BMC, bone mineral content. CTX, C-terminal cross-linked telopeptide; N, number. P1NP, serum procollagen type I N-propeptide. S.d., standard deviation.
**Footnotes.** 1, adjusted for baseline value and school of attendance. 2, deseasonalised values. 3, median calcium intake 466 mg/day.

### FRACTURES

17 participants reported 17 fractures during follow-up (11 upper limb, 4 lower limb and 2 at another anatomical site; Table S3, Supplementary Material). Allocation to vitamin D vs. placebo did not influence the proportion of participants reporting one or more fractures (adjusted odds ratio [aOR] 0.70, 95% CI 0.27 to 1.85, P=0.48; Table 4). Sub-group analyses evaluating the effects of the intervention by sex, baseline serum 25(OH)D_3_ concentration and calcium intake revealed no evidence of effect modification (P values for interaction ≥0.77 where calculable). Similarly null results were obtained for fractures reported as being X-ray-confirmed and for those reported as being treated with a plaster cast (Tables S4 and S5, Supplementary Material).

**Table 4.**
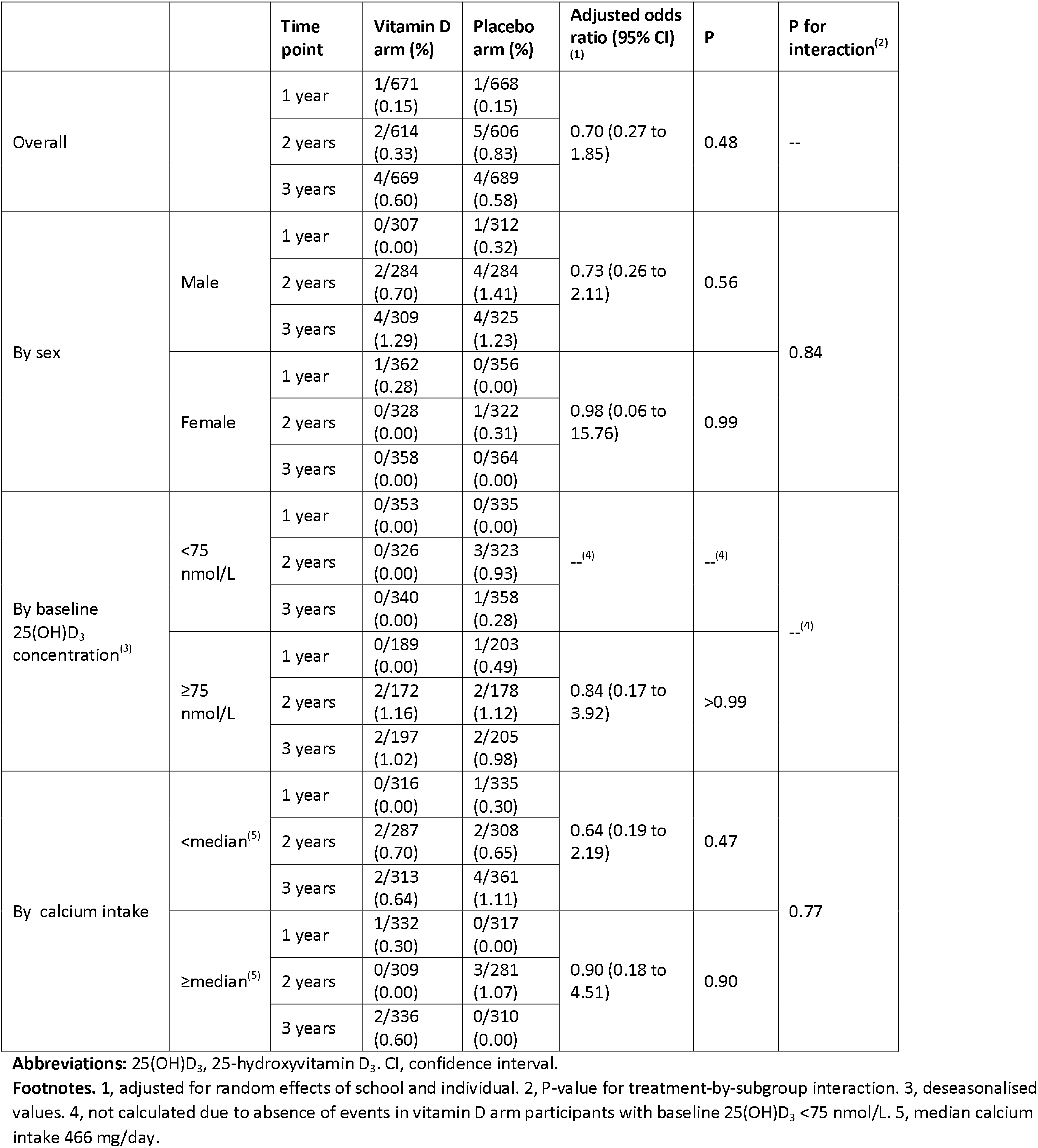
Proportion reporting one or more fractures by follow-up time point and allocation, overall and by sub-group

|  |  | Time point | Vitamin D arm (%) | Placebo arm (%) | Adjusted odds ratio (95% CI)<br>(1) | P | P for interaction<sup>(2)</sup> |
| --- | --- | --- | --- | --- | --- | --- | --- |
| Overall |  | 1 year | 1/671<br>(0.15) | 1/668<br>(0.15) | 0.70 (0.27 to 1.85) | 0.48 | -- |
|  |  | 2 years | 2/614<br>(0.33) | 5/606<br>(0.83) |  |  |  |
|  |  | 3 years | 4/669<br>(0.60) | 4/689<br>(0.58) |  |  |  |
| By sex | Male | 1 year | 0/307<br>(0.00) | 1/312<br>(0.32) | 0.73 (0.26 to 2.11) | 0.56 | 0.84 |
|  |  | 2 years | 2/284<br>(0.70) | 4/284<br>(1.41) |  |  |  |
|  |  | 3 years | 4/309<br>(1.29) | 4/325<br>(1.23) |  |  |  |
|  | Female | 1 year | 1/362<br>(0.28) | 0/356<br>(0.00) | 0.98 (0.06 to 15.76) | 0.99 |  |
|  |  | 2 years | 0/328<br>(0.00) | 1/322<br>(0.31) |  |  |  |
|  |  | 3 years | 0/358<br>(0.00) | 0/364<br>(0.00) |  |  |  |
| By baseline 25(OH)D<sub>3</sub> concentration<sup>(3)</sup> | <75 nmol/L | 1 year | 0/353<br>(0.00) | 0/335<br>(0.00) | --<sup>(4)</sup> | --<sup>(4)</sup> | --<sup>(4)</sup> |
|  |  | 2 years | 0/326<br>(0.00) | 3/323<br>(0.93) |  |  |  |
|  |  | 3 years | 0/340<br>(0.00) | 1/358<br>(0.28) |  |  |  |
|  | ≥75 nmol/L | 1 year | 0/189<br>(0.00) | 1/203<br>(0.49) | 0.84 (0.17 to 3.92) | >0.99 |  |
|  |  | 2 years | 2/172<br>(1.16) | 2/178<br>(1.12) |  |  |  |
|  |  | 3 years | 2/197<br>(1.02) | 2/205<br>(0.98) |  |  |  |
| By calcium intake | <median<sup>(5)</sup> | 1 year | 0/316<br>(0.00) | 1/335<br>(0.30) | 0.64 (0.19 to 2.19) | 0.47 | 0.77 |
|  |  | 2 years | 2/287<br>(0.70) | 2/308<br>(0.65) |  |  |  |
|  |  | 3 years | 2/313<br>(0.64) | 4/361<br>(1.11) |  |  |  |
|  | ≥median<sup>(5)</sup> | 1 year | 1/332<br>(0.30) | 0/317<br>(0.00) | 0.90 (0.18 to 4.51) | 0.90 |  |
|  |  | 2 years | 0/309<br>(0.00) | 3/281<br>(1.07) |  |  |  |
|  |  | 3 years | 2/336<br>(0.60) | 0/310<br>(0.00) |  |  |  |

**Abbreviations:** 25(OH)D<sub>3</sub>, 25-hydroxyvitamin D<sub>3</sub>. CI, confidence interval.
**Footnotes.** 1, adjusted for random effects of school and individual. 2, P-value for treatment-by-subgroup interaction. 3, deseasonalised values. 4, not calculated due to absence of events in vitamin D arm participants with baseline 25(OH)D<sub>3</sub> <75 nmol/L. 5, median calcium intake 466 mg/day.

### ADVERSE EVENTS

Incidence of adverse events by trial arm has been reported elsewhere.^15^ No serious events arising in the trial were adjudged related to administration of vitamin D or placebo.

## DISCUSSION

We report findings of the first RCT to investigate effects of vitamin D supplementation on BMC in children of Black African ancestry. Vitamin D insufficiency (25[OH]D 50-74.9 nmol/L) was common at baseline, and weekly oral administration of 10,000 IU vitamin D_3_ for 3 years was effective in suppressing serum PTH concentrations and elevating 25(OH)D concentrations above the 75 nmol/L threshold. However, these biochemical effects were not associated with changes in BMC or serum concentrations of bone turnover markers in the study population as a whole. No evidence for effect modification by baseline vitamin D status, sex or calcium intake was found. Neither was any effect of the intervention seen on incidence of fractures.

Null results of our trial for BMC outcomes contrast with positive findings from other studies that have investigated effects of higher-dose vitamin D on BMC. Fuleihan and colleagues reported that weekly administration of 14,000 IU vitamin D for one year increased bone area and total hip BMC in HIV-uninfected girls aged 10-17 years in Lebanon; sub-group analysis revealed that these changes were restricted to pre-menarcheal participants.^7^ Meta-analysis of RCT investigating effects of higher-dose vitamin D (1600 to 4000 IU/day) in HIV-infected adolescents and young adults living in the USA^8^ and Thailand^9^ also revealed vitamin D-induced increases in total BMC.^6^ Differences in outcomes between these studies vs. our own might reflect the relatively high baseline vitamin D status among participants in our study. However, results from our linked trial in Mongolian schoolchildren, ^19^ whose baseline vitamin D status was much lower than in the current study, were also null for BMC outcomes. Other potential explanations for our null findings for BMC outcomes include participants’ low calcium intakes, and (for comparison with results from USA/Thailand) the fact that our participants were not HIV-infected or taking anti-retroviral therapy - both factors that may modify effects of vitamin D supplements on BMC.^20^

Our study has several strengths. The placebo-controlled RCT design minimises potential for observer bias and confounding to operate. Administration of vitamin D supplements was sustained (3-year duration) and directly observed during term-time,and the dose administered was sufficient to elevate serum 25(OH)D concentrations into the physiological range (75-200 nmol/L) and to suppress PTH concentrations. We employed DXA, the gold standard methodology, to measure BMC, and complemented it with measurement of markers of bone formation and resorption. The bone sub-study was large, and loss to follow-up was lower than anticipated in the power calculation: accordingly, we were well powered to detect even modest effects of the intervention on BMC. Participants were less than 12 years old at enrolment, and were therefore exposed to the intervention before the period of peak bone mineral mass accretion, thereby maximising potential for the intervention to impact BMC and fracture risk. External validity was maximised by inclusion of both males and females.

Our study also has some limitations. Very few fractures were reported, which limited our power to detect an effect of the intervention on this outcome. Low fracture risk among participants in the current trial contrasts with the much higher event rate seen in our linked trial in Mongolia,^19^ consistent with reports that age-standardised fracture incidence in Southern Africa is among the lowest globally.^21^ Genotyping was not performed, so we were unable to test whether polymorphisms in the vitamin D receptor modified the effect of vitamin D supplementation on bone mineralisation, as has been reported by others.^22^ Vitamin D deficiency (25[OH]D <50 nmol/L) was uncommon at baseline, so our findings are not generalisable to populations with very low baseline vitamin D status; however, vitamin D insufficiency (25[OH]D 50-74.9 nmol/L) was common, and suppression of serum PTH concentrations among participants irrespective of their baseline vitamin D status reveals evidence of secondary hyperparathyroidism, reflecting the fact that many participants were not vitamin D-replete at baseline. We only investigated one vitamin D dosing regimen, without concomitant administration of calcium; our findings cannot therefore shed light on the question of whether higher or lower doses of vitamin D, with or without additional calcium supplementation, may impact BMC. Our estimates of calcium intake are approximate, due to the lack of food frequency questionnaires that have been validated for quantitation of calcium intake in the study population.

In conclusion, we report that oral administration of vitamin D at a dose of 10,000 IU/week for 3 years was effective in elevating vitamn D status and suppressing serum PTH concentrations in HIV-uninfected Black South African schoolchildren aged 10-11 years at baseline. However, these effects were not associated with changes in BMC or serum concentrations of bone turnover markers.

## Data Availability

Anonymised data may be requested from the corresponding author to be shared subject to terms of research ethics committee approval.

## CONTRIBUTORS

ARM conceived the study. KM, LKM, AKC, JN, CC, NCH, RLH, RJW, LGB and ARM contributed to study design and protocol development. KM led on trial implementation, with support from JS, CD, DAJ, JN, LGB and ARM. LKM and AEM oversaw performance of DXA scans. AvG advised on estimation of calcium intake. JCYT and WDF performed and supervised the conduct of biochemical assays. NW, RLH and ARM drafted the statistical analysis plan. DAJ, KM, JS, NW and CD managed data. NW, KM and JS accessed, verified and analysed the data underlying the study. ARM wrote the first draft of the trial report. All authors made substantive comments thereon and approved the final version for submission.

## ACKNOWLEDGEMENTS

This research was funded by the UK Medical Research Council (refs MR/R023050/1 and MR/M026639/1, both awarded to ARM). RJW was supported by Wellcome (104803, 203135). He also received support from the Francis Crick Institute which is funded by Cancer Research UK (FC2112), the UK Medical Research Council (FC2112) and Wellcome (FC2112). We thank all the children who participated in the trial, and their parents / guardians; members of the Independent Data Monitoring Committee (Prof Guy Thwaites, Oxford University Clinical Research Unit, Ho Chi Minh City, Vietnam [Chair]; Prof John Pettifor, University of the Witwatersrand, Johannesburg, South Africa; and Prof Sarah Walker, MRC Clinical Trials Unit, London, UK); and members of the Trial Steering Committee (Prof Beate Kampmann, London School of Hygiene and Tropical Medicine, London, UK [Chair]; Prof Ashraf Coovadia, University of the Witwatersrand, Johannesburg, South Africa; Dr Karen Jennings, City Health, Cape Town, South Africa; and Dr Guy de Bruyn, Sanofi Pasteur, Swiftwater PA USA). For the purposes of open access the author has applied a CC-BY public copyright to any author-accepted manuscript arising from this submission.

