## Supplementary Appendix for "Influence of vitamin D supplementation on bone mineral content, bone turnover markers and fracture risk in South African schoolchildren: multicentre double-blind randomised placebo-controlled trial (ViDiKids)"

**Supplementary Material**

### **Study Personnel**

**Chief Investigator:** Adrian R. Martineau (Queen Mary University of London)

**Principal Investigator:** Keren Middelkoop (University of Cape Town)

**Co-investigators:** Justine Stewart (University of Cape Town), David A. Jolliffe (Queen Mary University of London), Anna K Coussens (University of Cape Town), James Nuttall (Red Cross Children’s Hospital), Geeta Trilok Kumar (University of Delhi), Suzanne Filteau (London School of Hygiene and Tropical Medicine), Robert J Wilkinson (University of Cape Town), Linda-Gail Bekker (University of Cape Town)

**Trial Steering Committee:** Beate Kampmann (London School of Hygiene and Tropical Medicine), Ashraf Coovadia (University of the Witwatersrand), Guy de Bruyn (Sanofi Pasteur), Robin Dyers (Western Cape Government: Health), Karen Jennings (City of Cape Town Health Department)

**Data Monitoring Committee:** Guy Thwaites (Oxford University), John Pettifor (University of the Witwatersrand), Sarah Walker (MRC Clinical Trials Unit)

**Statisticians:** Neil Walker (Queen Mary University of London); Richard Hooper (Queen Mary University of London)

**Study coordinators:** Avril Masters, Venesia Du Toit, Carmen Delport (all University of Cape Town)

**Biochemistry laboratory team:** Jonathan CY Tang, William D Fraser (both University of East Anglia)

**QFT-Plus laboratory team:** Peter Meewes, Heloise Blignaut, Sarah Kellow-Webb, Lelo Mdluli, Jabu Khumalo, Lameez Appolis, Dineo Mathabela, Khumbelo Nekhumbe (all at the Bio Analytical Research Corporation, South Africa)

**Data management team:** Ashleigh Barnes, Ryan Johnson (both University of Cape Town)

**Study doctors:** Gregory Skinner, Kim Hall, Llewellyn Fleurs (all University of Cape Town)

**Study Nurses:** Veronica Bam, Phozie Daka, Petra Frances De Lange, Daleen Griessel, Nicola Kelly, Ntsikelelo Khuselo, Nelsiwe Kunene, Phila Mawu, Meshack Monakali, Sibonile Mqulwana, Odwa Ntola, Ncumisa Nzenze, Tebogo Rammala, Mathemba Siyoko, Phoebe Tshangela, Zukiswa Xabanisa, Jennifer Gelant, Xolani Gxako, Ferial Mahed (all University of Cape Town)

**Study Pharmacy team**: Edgar Barlow, Mary Chidanyika, Samantha Geduld, Fouche Geesje, Khadijah Gool, Ridley Howard, Amina Ismail, Julio Muller, Junaid Rawoot (all University of Cape Town)

**Logistics team:** Cynthia John, Pierre Kamunga, Marco Onkruid (all University of Cape Town)

**Field team:** Pearl Boks, Funeka Busakwe, Veliswa Cebisa, Vuyokazi Dinga, Tania Dinile, Dumani Domo, Nomakhosazana Gantsho, Bernadette Johnson, Xolani Madesi, Buyisile Mahlanza, Noluvuyo Mamkeli, Morris Manuel, Masibulele Matshoba, Nozicelo Mbiza, Nthuliseng Mohlafuno, Bongiwe Mtamzeli, Ndiseka Nashwa, Sibusiso Nduna, Thobile Nelani, Sihle Nqina, Melinda Onverwacht, Ntombenkosi Sipondo, Fikile Siqele , Babalwa Solinjani, Nonzaliseko Tempile, Nokukhanya Tiyane, Sindisiwe Tono, Bulelwa Zono (all University of Cape Town)

**Monitoring:** Catherine Lund, Robert Voogt (both from OnQ Research [Pty] Ltd)

### **Supplementary Methods**

#### ***Randomisation and blinding***

Eligible participants were individually randomised to receive a weekly capsule containing vitamin D_3_ or placebo for three years, with a one-to-one allocation ratio. Randomisation was stratified by school of attendance (a potential predictor of risk of QFT conversion) as follows. Prior to the start of recruitment, Mrs Claire Chan (Independent statistician, Pragmatic Clinical Trials Unit, Queen Mary University of London) prepared two school randomisation lists, each comprising 20 pairs of 2-letter randomisation codes, with each pair allocated to a single school identifier (e.g. School 1 was allocated codes ‘AC’ and ‘MO’, School 2 was allocated codes ‘BD’ and ‘NP’). One 2-letter randomisation code within each pair was then randomly assigned to the vitamin D arm of the trial, and the other was assigned to the placebo arm, using a computer-generated random sequence (e.g. ‘AC’ and ‘BD’ were assigned to ‘vitamin D’, ‘MO’ and ‘NP’ were assigned to placebo). Mrs Chan also prepared 40 separate participant randomisation lists (one for each potential participating school). Each of these participant randomisation lists comprised 999 5-digit numbers, each consisting of a 2-digit school identifier from 01 to 25 that was constant for, and unique to, each list, followed by a 3-digit participant identifier from 001 to 999 (e.g. 01-001, 01-002, 02-001, 02-002). These 5-digit numbers were then randomly assigned to one or other of the 2-letter randomisation codes allocated to that school in blocks of ten, using a computer-generated random sequence (e.g. for School 1, sequence was 01-001-MO, 01-002-AC, etc.; for School 2, sequence was 02-001-BD, 02-002-BD, etc.).

Active and placebo capsules were shipped from Tishcon Corporation (Westbury, NY, USA) to Lekoko Pharmaceutical Management Consultancy (Johannesburg, South Africa) in boxes that were labelled ‘vitamin D’ or ‘placebo’ according to their contents. On arrival, these capsules were packed into bottles, each containing either 150 vitamin D capsules or 150 placebo capsules. The school randomisation list was then used to label these bottles with 2-letter randomisation codes according to their contents (i.e. bottles containing vitamin D capsules were labelled with ‘AC’, ‘BD’ or another 2-letter code assigned to the vitamin D arm of the trial, while bottles containing placebo capsules were labelled ‘MO’, ‘NP’ or another 2-letter code assigned to the placebo arm of the trial) until sufficient capsules for 23 schools had been bottled and labelled. Bottling and labelling was performed under the supervision of Mr Bobby Hamman (Lekoko PMC), with independent monitoring performed by staff from OnQ Contract Research Organisation; none of these individuals was involved with data collection. Children screened at each school were assigned consecutive 5-digit numbers at enrolment by study field workers, and if they were subsequently found to be eligible for randomisation (i.e. if their baseline QFT-Plus result was negative) then the participant randomisation list for their school of attendance was used to determine their allocation, i.e. they received study medication from bottles labelled with the 2-letter code linked to their 5-digit ID in the participant randomization list for the duration of the trial. For example, participant 01-001 would receive study medication labelled ‘MO’ (i.e. placebo), 01-002 would receive medication labelled ‘AC’ (i.e. vitamin D), 02-001 would receive medication labelled ‘BD’ (vitamin D), and 02-002 would receive medication labelled ‘BD’ (vitamin D) throughout the trial. Copies of the school randomisation list were held by Mrs Chan and by members of the DSMB. Neither participants nor trial staff had access to it, and treatment allocation was concealed from participants, care providers and all trial staff (including senior investigators and those assessing outcomes) so that the double-blind was maintained. The school randomisation lists were made available to Dr Neil Walker (trial statistician) following completion of the trial, who used them to un-blind allocation and to analyse the trial: he was not therefore masked to group assignment during statistical analysis.

#### ***Calculation of calcium intake***

Dietary calcium intake was calculated on the basis of parental reports of participants’ intake of oily fish, eggs, margarine, liver, red meat, fresh milk, sour milk, cheese, yoghurt, beans and green vegetables in the month prior to their enrolment, captured using an electronic report form (Figure S1). Calcium content per portion for each item was calculated by multiplying average portion size for children aged 6-11 years, derived using South African food guide unit sizes^1^ by the typical calcium content of each food, as detailed in the South African Food Composition Database 2021.^2^ For each calcium source, this estimate of calcium content in a typical portion was multiplied by the weekly number of portions consumed. The calcium intake from each source was then summed and divided by 7 to yield daily calcium intake (range 57 to 1467 mg/day). For sub-group analyses, calcium intake was dichotomised according to whether the value was below vs. above or equal to the median value of 466 mg/day.

### **Table S1**. Values for calcium content and portion size of calcium-containing foods used to calculate calcium intake.

|  |  | Calcium content (mg per gram)^(1)^ | Average portion size for child aged 6-11 years (grams or ml)^(2)^ |
| --- | --- | --- | --- |
| Oily fish | Pilchard (canned) | 3.6 | 125 |
|  | Sardines (canned) | 3.1 | 125 |
|  | Snoek | 0.3 | 120 |
|  | Bokkoms (salted) | 0.7 | 40 |
|  | Tuna (canned) | 0.1 | 150 |
|  | Mackerel | 0.2 | 120 |
|  | Salmon (canned) | 2.15 | 125 |
| Eggs |  | 0.4 | 100 |
| Margarine |  | 0.1 | 7 |
| Liver (beef, lamb, chicken or pork) |  | 0.1 | 100 |
| Red meat (beef or lamb) |  | 0.2 | 80 |
| Fresh milk |  | 1.1 | 200 |
| Sour milk |  | 1.6 | 200 |
| Cheese |  | 8.1 | 40 |
| Yoghurt |  | 1.4 | 100 |
| Baked beans or other beans, e.g. kidney |  | 0.3 | 75 |
| Green vegetables, e.g. spinach, broccoli |  | 0.6 | 80-100 |

Footnotes: **1,** estimates from the South African Food Composition Database (2021).^2^ **2,** estimate from proposed South African Food Guide Unit Sizes.^1^

### **Table S2.** Corrected^(1)^ end-trial bone mineral content at the whole body minus head and lumbar spine sites by allocation: overall and by sub-groups

|  |  | **Vitamin D arm: mean value, g (s.d.) [N]** | **Placebo arm: mean value, g (s.d.) [N]** | **Adjusted mean difference (95% CI)^(2)^** | **P** | **P for interaction** |
| --- | --- | --- | --- | --- | --- | --- |
| **Whole body minus head** | | | | | | |
| Overall |  | 1112.9 (255.3) [202] | 1071.5 (221.9) [189] | 6.4 (-4.2 to 17.0) | 0.24 | -- |
| By sex | Male | 1102.3 (268.5) [97] | 1031.4 (225.4) [89] | 8.1 (-7.5 to 23.8) | 0.31 | 0.60 |
|  | Female | 1122.7 (243.4) [105] | 1107.2 (213.5) [100] | 2.9 (-10.8 to 16.7) | 0.68 |  |
| By baseline 25(OH)D_3_ concentration^(3)^ | <75 nmol/L | 1137.0 (270.8) [103] | 1099.1 (224.2) [96] | 8.0 (-6.6 to 22.6) | 0.28 | 0.99 |
|  | ≥75 nmol/L | 1119.3 (269.6) [60] | 1058.4 (215.9) [54] | 8.7 (-12.9 to 30.2) | 0.43 |  |
| By calcium intake | <median^(4)^ | 1111.3 (249.5) [94] | 1057.8 (212.9) [97] | 11.4 (-3.8 to 26.6) | 0.14 | 0.30 |
|  | ≥median^(4)^ | 1120.7 (263.6) [102] | 1094.0 (228.9) [89] | 1.3 (-13.7 to 16.2) | 0.87 |  |
| **Lumbar spine** | | | | | | |
| Overall |  | 36.2 (10.1) [202] | 34.2 (8.0) [189] | 0.3 (-0.7 to 1.2) | 0.58 | -- |
| By sex | Male | 33.2 (9.7) [97] | 31.1 (6.8) [89] | 0.3 (-0.8 to 1.3) | 0.64 | 0.92 |
|  | Female | 39.0 (9.7) [105] | 37.0 (8.0) [100] | 0.3 (-0.7 to 1.3) | 0.56 |  |
| By baseline 25(OH)D_3_ concentration^(3)^ | <75 nmol/L | 37.5 (10.7) [103] | 35.4 (8.4) [96] | 0.7 (-0.6 to 1.9) | 0.29 | 0.85 |
|  | ≥75 nmol/L | 36.2 (10.5) [60] | 33.1 (6.9) [54] | 0.3 (-1.6 to 2.1) | 0.76 |  |
| By calcium intake | <median^(4)^ | 35.5(9.9) [94] | 33.9 (7.8) [97] | 0.3 (-1.0 to 1.5) | 0.70 | 0.97 |
|  | ≥median^(4)^ | 36.9 (10.2) [102] | 34.8 (8.2) [89] | 0.1 (-1.2 to 1.5) | 0.83 |  |

**Abbreviations:** 25(OH)D_3_, 25-hydroxyvitamin D_3_. BMC, bone mineral content. CI, confidence interval. S.d., standard deviation. N, number.

**Footnotes.** 1, i.e. with volumetric correction and correction for bone area, height and weight. **2,** adjusted for baseline value and school of attendance. 3, deseasonalised values. 4, median calcium intake 466 mg/day.

### **Table S3.** Fracture incidence by anatomical site, overall and by allocation.

|  |  | Number of participants reporting ≥1 fracture | | | Number of fractures reported | | |
| --- | --- | --- | --- | --- | --- | --- | --- |
|  |  | Overall | Vitamin D arm | Placebo arm | Overall | Vitamin D arm | Placebo arm |
| Anatomical site | Lower limb | 4 | 2 | 2 | 4 | 2 | 2 |
|  | Upper limb | 11 | 5 | 6 | 11 | 5 | 6 |
|  | Other | 2 | 0 | 2 | 2 | 0 | 2 |
| **Total** |  | 17 | 7 | 10 | 17 | 7 | 10 |

### **Table S4.** Proportion reporting one or more X-ray-confirmed fractures by follow-up time point and allocation, overall and by sub-group

|  |  | **Time point** | **Vitamin D arm (%)** | **Placebo arm (%)** | **Adjusted odds ratio (95% CI) ^(1)^** | **P** | **P for interaction^(2)^** |
| --- | --- | --- | --- | --- | --- | --- | --- |
| Overall |  | 1 year | 1/671 (0.15) | 1/668 (0.15) | 0.94 (0.29 to 3.09) | 0.92 | -- |
|  |  | 2 years | 2/614 (0.33) | 3/604 (0.50) |  |  |  |
|  |  | 3 years | 5/669 (0.75) | 4/689 (0.58) |  |  |  |
| By sex | Male | 1 year | 0/307 (0.00) | 1/312 (0.32) | 1.03 (0.29 to 3.74) | 0.96 | 0.98 |
|  |  | 2 years | 2/284 (0.70) | 2/282 (0.71) |  |  |  |
|  |  | 3 years | 5/309 (1.62) | 4/325 (1.23) |  |  |  |
|  | Female | 1 year | 1/362 (0.28) | 0/356 (0.00) | 0.98 (0.06 to 15.76) | 0.99 |  |
|  |  | 2 years | 0/328 (0.00) | 1/322 (0.31) |  |  |  |
|  |  | 3 years | 0/358 (0.00) | 0/364 (0.00) |  |  |  |
| By baseline 25(OH)D_3_ concentration^(3)^ | <75 nmol/L | 1 year | 0/353 (0.00) | 0/335 (0.00) | --^(4)^ | --^(4)^ | --^(4)^ |
|  |  | 2 years | 0/326 (0.00) | 2/322 (0.62) |  |  |  |
|  |  | 3 years | 0/340 (0.00) | 1/358 (0.28) |  |  |  |
|  | ≥75 nmol/L | 1 year | 0/189 (0.00) | 1/203 (0.49) | 1.05 (0.19 to 5.66 | >0.99 |  |
|  |  | 2 years | 2/172 (1.16) | 1/177 (0.56) |  |  |  |
|  |  | 3 years | 3/197 (1.52) | 2/205 (0.98) |  |  |  |
| By calcium intake | <median^(5)^ | 1 year | 0/316 (0.00) | 1/335 (0.30) | 0.68 (0.16 to 2.83) | 0.59 | 0.31 |
|  |  | 2 years | 2/287 (0.70) | 2/308 (0.65) |  |  |  |
|  |  | 3 years | 3/313 (0.96) | 4/361 (1.11) |  |  |  |
|  | ≥median^(5)^ | 1 year | 1/332 (0.30) | 0/317 (0.00) | 2.70 (0.28 to 26.29) | 0.39 |  |
|  |  | 2 years | 0/309 (0.00) | 1/279 (0.36) |  |  |  |
|  |  | 3 years | 2/336 (0.60) | 0/310 (0.00) |  |  |  |

**Abbreviations:** 25(OH)D_3_, 25-hydroxyvitamin D_3_. CI, confidence interval.

**Footnotes.** 1, adjusted for random effects of school and individual. 2, P-value for treatment-by-subgroup interaction. 3, deseasonalised values. 4, not calculated due to zero events in vitamin D arm participants with baseline 25(OH)D_3_ <75 nmol/L. 5, median calcium intake 466 mg/day.

### **Table S5.** Proportion reporting one or more plaster cast-treated fractures by follow-up time point and allocation, overall and by sub-group

|  |  | **Time point** | **Vitamin D arm (%)** | **Placebo arm (%)** | **Adjusted odds ratio (95% CI) ^(1)^** | **P** | **P for interaction^(2)^** |
| --- | --- | --- | --- | --- | --- | --- | --- |
| Overall |  | 1 year | 1/671 (0.15) | 1/668 (0.15) | 0.76 (0.22 to 2.67) | 0.67 | -- |
|  |  | 2 years | 2/614 (0.33) | 3/605 (0.50) |  |  |  |
|  |  | 3 years | 4/669 (0.60) | 4/689 (0.58) |  |  |  |
| By sex | Male | 1 year | 0/307 (0.00) | 1/312 (0.32) | 0.79 (0.21 to 3.02) | 0.74 | 0.86 |
|  |  | 2 years | 2/284 (0.70) | 2/283 (0.71) |  |  |  |
|  |  | 3 years | 4/309 (1.29) | 4/325 (1.23) |  |  |  |
|  | Female | 1 year | 1/362 (0.28) | 0/356 (0.00) | 0.98 (0.06 to 15.73) | 0.99 |  |
|  |  | 2 years | 0/328 (0.00) | 1/322 (0.31) |  |  |  |
|  |  | 3 years | 0/358 (0.00) | 0/364 (0.00) |  |  |  |
| By baseline 25(OH)D_3_ concentration^(3)^ | <75 nmol/L | 1 year | 0/353 (0.00) | 0/335 (0.00) | --^(4)^ | --^(4)^ | --^(4)^ |
|  |  | 2 years | 0/326 (0.00) | 1/322 (0.31) |  |  |  |
|  |  | 3 years | 0/340 (0.00) | 1/358 (0.28) |  |  |  |
|  | ≥75 nmol/L | 1 year | 0/189 (0.00) | 1/203 (0.49) | 0.84 (0.17 to 3.92) | 0.50 |  |
|  |  | 2 years | 2/172 (1.16) | 2/178 (1.12) |  |  |  |
|  |  | 3 years | 3/197 (1.52) | 2/205 (0.98) |  |  |  |
| By calcium intake | <median^(5)^ | 1 year | 0/316 (0.00) | 1/335 (0.30) | 0.81 (0.18 to 3.58)^(6)^ | 0.78 | 0.87 |
|  |  | 2 years | 2/287 (0.70) | 1/308 (0.32) |  |  |  |
|  |  | 3 years | 3/313 (0.96) | 4/361 (1.11) |  |  |  |
|  | ≥median^(5)^ | 1 year | 1/332 (0.30) | 0/317 (0.00) | 0.92 (0.13 to 6.56)^(6)^ | 0.94 |  |
|  |  | 2 years | 0/309 (0.00) | 2/280 (0.71) |  |  |  |
|  |  | 3 years | 1/336 (0.30) | 0/310 (0.00) |  |  |  |

**Abbreviations:** 25(OH)D_3_, 25-hydroxyvitamin D_3_. CI, confidence interval.

**Footnotes.** 1, adjusted for random effects of school and individual. 2, P-value for treatment-by-subgroup interaction. 3, deseasonalised values. 4, not calculated due to zero events in vitamin D arm participants with baseline 25(OH)D_3_ <75 nmol/L. 5, median calcium intake 466 mg/day. 6, not adjusted for random effect of school due to non-convergence.

### **Figure S1.** Case report form capturing details of dietary intake of foods containing vitamin D and/or calcium in the previous month

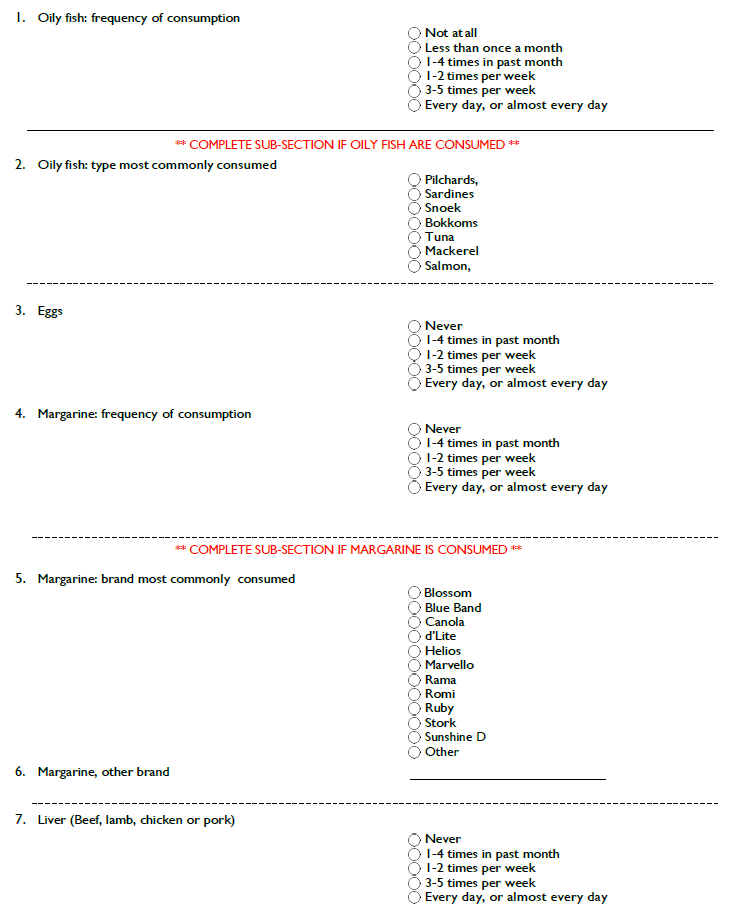

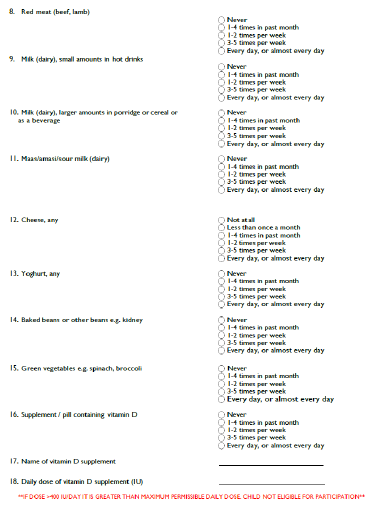

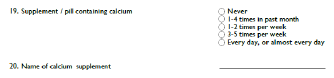

### **Figure S2.** Case report form capturing details of incident fractures in previous year

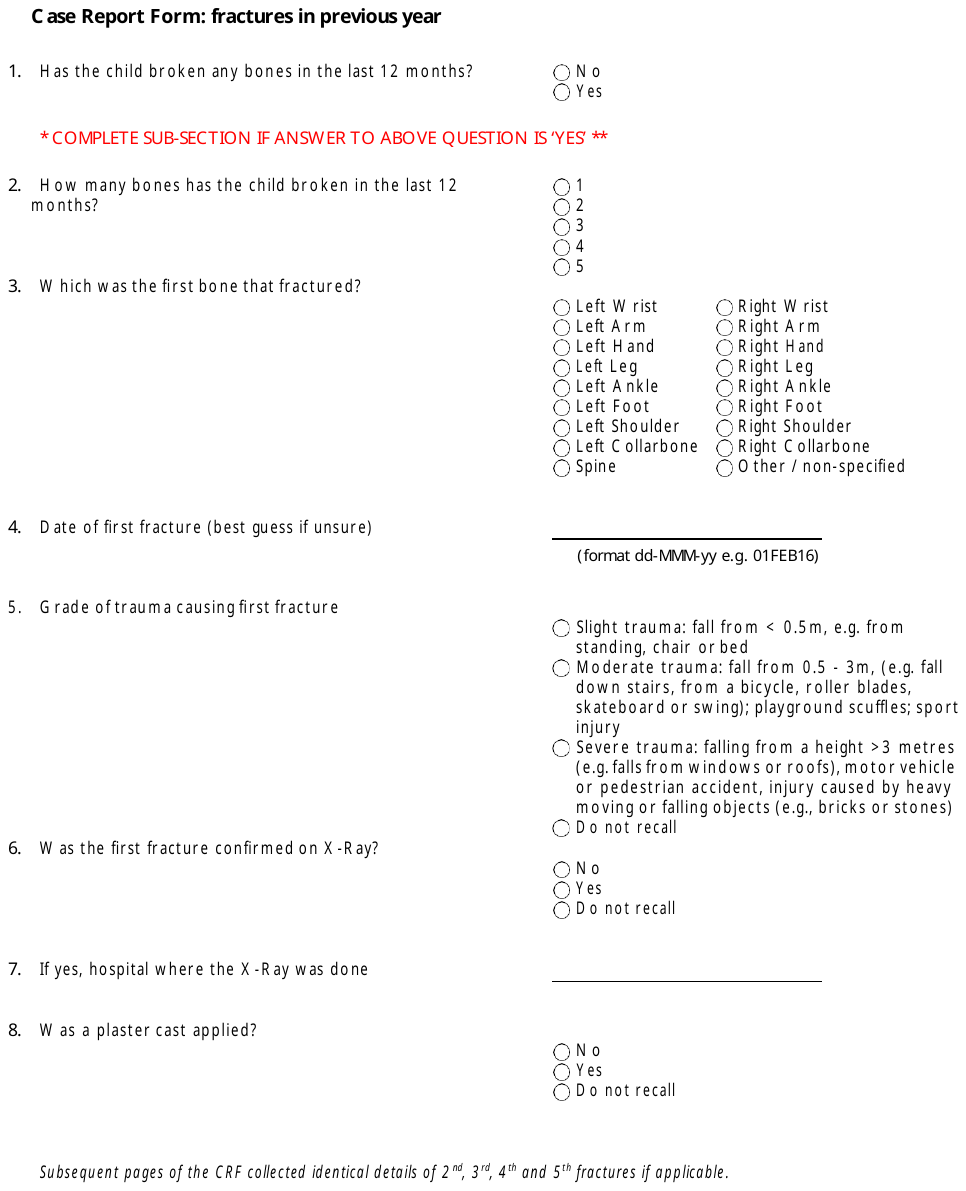

2.	South African Medical Research Council. South African Food Database System (SAFOODS). Food Composition Database. Parow Valley, Cape Town; 2021.
